# Design and Rationale of the My Heart Counts Cardiovascular Health Study: a Large-Scale, Fully Digital Biobank, and Randomized Trial of Large Language Model-Driven Coaching of Physical Activity

**DOI:** 10.64898/2026.03.02.26347447

**Authors:** Paul Schmiedmayer, Anders Johnson, Narayan Schuetz, Lukas Kollmer, Paul Goldschmidt, Juan Delgado-SanMartin, Kelly W Zhang, Sriya Mantena, Alex Tolas, Samuel Montalvo, Mariana Ramirez-Posada, Jack W. O’Sullivan, Marily Oppezzo, Abby C King, Fatima Rodriguez, Euan Ashley, Allan Lawrie, Daniel Seung Kim

**Author notes:** These authors contributed equally to the supervision of this work (Euan Ashley, Allan Lawrie, Daniel Seung Kim). <u>Corresponding authors:</u> Euan A. Ashley, MB, ChB, DPhil; Allan Lawrie, PhD; Daniel Seung Kim, MD, PhD, MPH.

## Abstract

**Background:** Cardiovascular disease remains the leading cause of global morbidity and mortality. The original *My Heart Counts* smartphone application demonstrated the feasibility of large-scale, fully digital recruitment and trial conduct, but was limited by platform exclusivity and the need for human experts to create text-based behavioral interventions.

**Methods:** The next-generation *My Heart Counts* smartphone application is a prospective, observational cohort study with an embedded randomized crossover trial, evaluating personalized text-based coaching prompts, available in both English and Spanish. All study and trial operations will be conducted via the *My Heart Counts* smartphone application, re-designed using the open-source Stanford Spezi framework to support iOS, with a planned Android release in 2027. The target enrollment is N=15,000 adults across the United States and United Kingdom. The study establishes a comprehensive digital biobank by synthesizing passive mobile health data (steps, flights climbed, heart rate, sleep, workouts), raw sensor data (e.g., accelerometry), longitudinal clinical surveys, active tasks (6-minute walk test and 12-minute Cooper run test), electrocardiograms (ECG), and electronic health record (EHR) data integrated via HL7 FHIR protocols. The embedded trial evaluates the effect of text-based coaching prompts generated by a large language model (LLM) grounded in the Transtheoretical Model of Change on daily physical activity, as compared to generic prompts.

**Planned Analysis:** The primary endpoint of the randomized crossover trial is change in daily step count between LLM-driven and generic text-based intervention arms, analyzed using mixed-effects models. Secondary endpoints include change in mean active minutes and calorie burn over each intervention week. Other analyses include the changes in submaximal (6-minute walk test) and maximal (Cooper 12-minute run test) cardiorespiratory fitness, changes to sensor-derived biomarkers (e.g., sleep quality, resting heart rate, and heart rate variability), and association of sensor-derived biomarkers with EHR-confirmed clinical outcomes.

**Conclusions:** By utilizing autonomous, LLM-driven coaching, modular software design, and cross-platform accessibility, our smartphone application-based study will provide a scalable model for inclusive and decentralized preventive care of patients with cardiovascular disease.

**Trial Status:** Recruitment commenced in March 2026 and is ongoing.

## BACKGROUND

Cardiovascular disease remains the leading cause of mortality worldwide, accounting for approximately 19 million deaths annually^1^. Physical inactivity represents one of the most significant modifiable risk factors for cardiovascular disease^2^. Yet despite compelling evidence for the benefits of regular physical activity, the majority of American adults and 45% of European adults fail to meet modest recommendations of at least 150 minutes of moderate-intensity physical activity per week^3,4^. As a result, persistent physical inactivity contributes substantially to the global burden of cardiovascular disease, diabetes, cancer, and premature morbidity and mortality^5^.

Moreover, patients often have a poor understanding of their cardiovascular disease risk^6^. This is associated with a lack of symptoms associated with many of the intermediate risk factors (i.e., hypertension, diabetes, and hyperlipidemia)^6,7^. Patient-facing frameworks have been introduced that reduce statistics and probabilities to more holistic checklists and scores, creating a more actionable roadmap to reducing cardiovascular disease risk^8,9^. This approach empowers patients and allows them to see how small, incremental changes to modifiable risk factors (e.g., physical activity) reduce their cardiovascular disease risk, transforming abstract medical risk into a more manageable, personal goal.

Additionally, traditional approaches to promoting physical activity, including in-person coaching and group-based interventions, face inherent scalability limitations^10^. Provider time constraints, geographic barriers (i.e., rural vs urban environments), and the substantial costs associated with personalized coaching restrict the reach of these interventions to a small fraction of those who would benefit^11^. In this context, the widespread adoption of smartphones and wearable devices presents an unprecedented opportunity to deliver behavioral interventions at scale^10^.

## RATIONALE

The original *My Heart Counts*, launched in 2015 as a smartphone application on iOS, demonstrated the potential for rapid, large-scale enrollment, capturing data from >40,000 participants in the first two weeks and ultimately engaging >100,000 users^12–17^. This platform enabled the first fully digital randomized trial using a smartphone application, and demonstrated the utility of personalized, text-based coaching on increasing short-term physical activity^18,19^.

While the first iteration of the *My Heart Counts Cardiovascular Health Study* provided numerous novel insights, it faced significant technical and structural constraints. These included platform exclusivity to iOS, a lack of linguistic diversity (requiring written English comprehension for participation), use of submaximal fitness assessments, and the requirement for human experts to create and tailor the personalized text-based coaching prompts used in randomized trials^10,20,21^. To address these gaps and pioneer the next generation of digital health research, we have redesigned the *My Heart Counts* smartphone application using the Stanford Spezi framework, an open-source ecosystem dedicated to modern, interoperable digital health applications^22^.

The redesigned *My Heart Counts* smartphone application will serve several purposes. First, it will operate as a comprehensive digital biobank for longitudinal cardiovascular health data, such as traditional risk factors like blood pressure, lipid and glucose levels, and weight, in addition to wearable-derived data such as heart rate variability, resting heart rate, physical activity and sleep time, and numerous other insights related to health. The application retrospectively collects available mobile health data up to 10 years before enrollment into the *My Heart Counts* study, allowing for pre-/post-diagnosis comparisons, where data is available^22^. Second, it will serve as the largest study to incorporate raw sensor data (e.g., accelerometry, photoplethysmography (PPG), time spent in ambient daylight, etc) collected in the raw representation and original sampling frequency by compatible wearable devices, e.g., using Apple SensorKit^23^. Third, it will introduce a patient-facing dashboard, summarizing their objective data and cumulative cardiovascular disease risk^8,9^. By seeing how their risk changes with modifiable risk factors (e.g., physical activity and sleep), we aim to motivate patients to make healthier choices day-by-day with the goal of reducing cardiovascular morbidity and mortality. Finally, the redesigned *My Heart Counts* smartphone application will serve as a digital platform and host for fully autonomous, randomized trials, which can be performed in numerous disease states (e.g., pulmonary arterial hypertension, type 2 diabetes, and heart failure) and in multiple languages (English and Spanish at launch, with plans for expansion to other languages in the future)^10,20,24^. The extensible software system is designed to support multiple sub-studies within the same mobile application, enabling additional randomized trials in addition to active tasks, questionnaires, educational content, and both retrospective/prospective data collection for cohorts enrolling through the *My Heart Counts* application^22^.

Central to this new software architecture is an embedded randomized crossover trial evaluating the efficacy of large language model (LLM)-driven physical activity coaching^10,25^. By utilizing customized LLMs grounded in behavioral psychology, these trials seek to overcome the limitations of static, human-crafted messaging used in previous digital health interventions^10,25^. Furthermore, the integration of electronic health record (EHR) data through standardized HL7 Fast Healthcare Interoperability Resources (FHIR) protocols bridges the gap between an individual’s wearable sensor data and clinical outcomes, creating a high-fidelity longitudinal record of a participant’s health journey and offering a visual interface for users to track and see how their cardiovascular risk can be modified through healthy behaviors^26^. This manuscript details the comprehensive design, methodology, and rationale for the next-generation smartphone application and its embedded trial, providing a blueprint for the future of precision cardiovascular medicine.

## METHODS

### Funding

Funding from the American Diabetes Association (ADA 7-25-INI-11), American Heart Association (25CDA1436622), Imperial BHF Research Excellence Award (4) (RE/24/130023), British Heart Foundation (RG/F/25/110167), National Institutes of Health (1L30HL170306-01 and 9L30DK144879-02), and Stanford Center for Digital Health were used to support the research and creation of this paper. The authors are solely responsible for the design and conduct of this study, all study analyses, the drafting and editing of this paper, and its final content.

### Study Design and Participant Population

The next-generation *My Heart Counts* smartphone application is designed as a prospective, observational cohort study with an embedded interventional trial. The study is managed by Stanford University and Imperial College London, and is conducted entirely in a virtual environment, with recruitment focused across the United States and United Kingdom. Future enrollment in other sites (e.g., the European Union and North/Central/South Americas) is planned.

**Figure 1.**
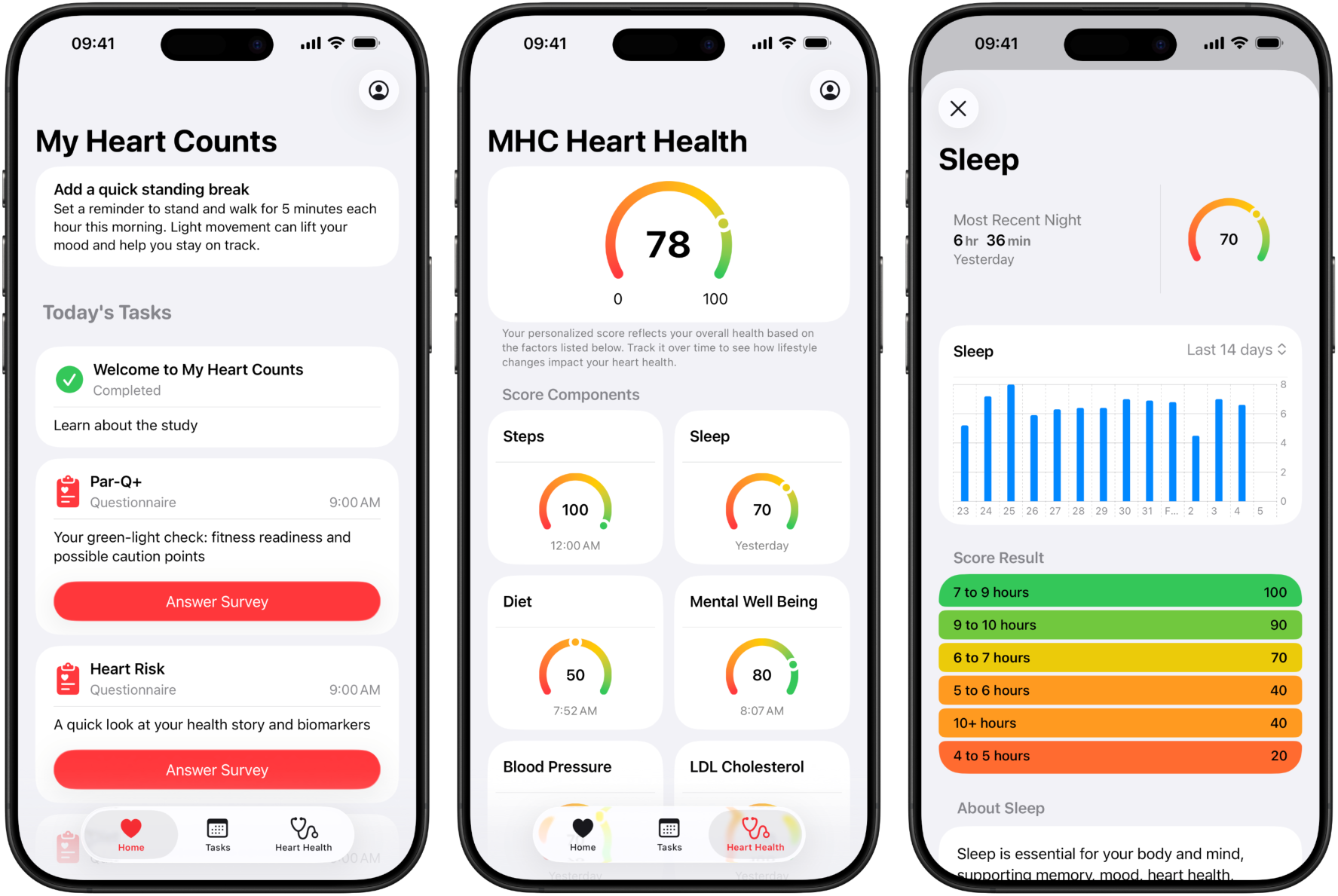
Overview of the user interface of the *My Heart Counts* smartphone application.

The study population comprises all adults aged 18 and older who are residents of either the United States or United Kingdom, and possess a compatible smartphone. To maximize inclusivity, the application is available in both English and Spanish (with other languages such as Chinese in development) and will be offered across iOS (available from March 2026) and Android devices (coming in 2027). The recruitment strategy explicitly aims to reach populations of varied age, racial, ethnic, socioeconomic and cardiovascular backgrounds^20,21,24^.

### Inclusion and Exclusion Criteria

Participants must be at least 18 years of age and able to read and understand the consent documentation, in either English or Spanish. We will make particular efforts to recruit individuals with cardiometabolic disease, including type 2 diabetes and obesity, and also those with rare cardiovascular diseases such as pulmonary arterial hypertension^15^. The exclusion criteria are minimal, but include the use of a shared Apple/Google Play ID, as this can lead to inaccuracies in the collection of smartphone sensor and wearable data. Furthermore, participants must be residents of either the United States or United Kingdom to align with current regulatory and institutional and/or ethics review board approvals.

### eConsent and Enrollment Process

Enrollment is facilitated through a streamlined, patient-centered eConsent process within the application^27^. This process uses icons and digestible text segments to explain the study’s purpose, risks, and benefits. A mandatory quiz assesses participants’ understanding of the consent, including that participation is voluntary and that they may withdraw at any time without negative consequences. After completing the quiz, participants provide an electronic signature, and a copy of the consent form is emailed to the participant and stored in the application’s “Account” tab for future reference. Participants are onboarded to the *My Heart Counts* smartphone application over the period of a week and asked to complete varied surveys (see **Table 1**) and complete a 6-minute walk test^14,15^ or Cooper 12-minute run test^28^ for assessment of their submaximal or maximal cardiorespiratory fitness, respectively.

**Table 1.** Overview of Task Schedule for 1-Week Onboarding.

| **Day** | **Tasks and Surveys** |
| --- | --- |
| **Day 1** | Welcome Informational<br>Demographics/Profile<br>PAR-Q+ (Screener before physical activity)<br>Heart Risk Factors<br>Diet Survey |
| **Day 2** | Survey of Mental Well-Being (WHO-5)<br>Disease state quality of life (for patients with angina, heart failure, or pulmonary hypertension)<br>Electrocardiogram (ECG) Active Task (if Apple Watch available) |
| **Day 3** | Sleep chronotype questionnaire<br>Questionnaire on fatigue levels |
| **Day 4** | Survey on self-reported activity/fitness levels |
| **Day 5** | Survey on symptoms of anxiety (GAD-7) |
| **Day 6** | Survey of perceived adequacy and processes involved in physical activity |
| **Day 7** | System usability survey (feedback on smartphone application design/use)<br>Information on the randomized crossover trial and option to opt-out |
| **Day 8** | Active task to complete either a submaximal (6-minute walk test) or maximal (Cooper 12-minute run test) cardiorespiratory fitness evaluation<br>Randomized crossover trial begins |

### Data Collection: Wearables, Surveys, and EHR Integration

The *My Heart Counts* smartphone application has a multi-layered approach to data collection, which synthesizes self-reported survey data with objective biometric sensors and EHR data. All questionnaires, data collected, and relevant metadata is transformed into uniform Health Level Seven (HL7) FHIR (Fast Healthcare Interoperability Resources) representations and uniform coding schemas^26^. This comprehensive data environment will allow for validation of digital biomarkers with traditional clinical markers.

#### Wearable and Sensor Data

The *My Heart Counts* smartphone application uses Apple HealthKit and SensorKit (on iOS) and Google Health Connect (on Android, from 2027) frameworks to passively capture high-frequency health data (see **Table 2**). We plan to extend the data collection to Google Fit (Fitbit), Withings, and OURA devices in 2027. This passive data stream provides a continuous view of the participant’s lifestyle that cannot be captured during episodic clinic visits. In addition to aggregated metrics, the application will collect raw triaxial accelerometry and gyroscope data during active tasks to facilitate advanced signal processing and development of novel motion algorithms. If participants grant permission to collect raw sensor data via SensorKit (e.g., photoplethysmography (PPG), unprocessed electrocardiogram (ECG) voltage measurements, accelerometer, barometer, ambient light exposure, mobility and device usage patterns, location and visit history, and more), the application will receive Apple-provided raw data exports from Apple Watches and iPhones after a platform-defined delay^23^.

**Table 2.** Passively collected metrics via Apple HealthKit or Google Health Connect.

| **Category** | **Metrics Collected** |
| --- | --- |
| **Physical Activity** | Daily steps, distance walking/running, flights climbed, active energy burned, active energy time, pedometer cadence. |
| **Gait and Mobility** | Walking speed, step length, walking asymmetry |
| **Vital Signs** | Heart rate (resting, walking), heart rate variability (SDNN), oxygen saturation, respiratory rate, wrist/skin temperature |
| **Cardiorespiratory** | Estimated maximal oxygen uptake (VO<sub>2</sub>max estimated by device), heart rate recovery, raw PPG waveform samples, raw single-lead ECG voltage traces |
| **Sleep** | Sleep analysis (total duration, deep/light/REM stages), sleep time consistency |
| **Symptoms** | High/low heart rate events, irregular heart rhythm events (e.g., Atrial Fibrillation burden), |
|  | ECG classifications |
| **Context and Environmental Signals** | Ambient light exposure, ambient pressure (barometer), environmental audio level summaries, accelerometer (raw 3-axis motion), device wear time (on-wrist detection), mobility patterns (visits and dwell time), device usage patterns (interaction summaries) |

#### Longitudinal Survey Instrumentation

The *My Heart Counts* smartphone application employs numerous surveys to capture health-related quality of life and behavioral determinants (see **Table 3**). All surveys are all compatible with Fast Healthcare Interoperability Resources (FHIR) standards, which enables structured, interoperable data exchange for patient-reported outcomes^26^.

**Table 3.** Longitudinal Surveys.

| **Survey Name** | **Population** | **Purpose** | **Frequency** |
| --- | --- | --- | --- |
| **Seattle Angina Questionnaire 7 (SAQ-7)<sup>29</sup>** | Coronary Artery Disease | Disease-specific HRQOL | Every 2 weeks (if symptomatic) |
| **Kansas City Cardiomyopathy Questionnaire 12 (KCCQ-12)<sup>30</sup>** | Heart failure | Disease-specific HRQOL | Every 2 weeks (if symptomatic) |
| **EmPHasis-10<sup>31</sup>** | Pulmonary Arterial Hypertension | Disease-specific HRQOL | Every 2 weeks (if symptomatic) |
| **Fatigue Survey** | Everyone | Effect of fatigue on HRQOL | Every 2 weeks (if symptomatic) |
| **WHO-5<sup>32</sup>** | Everyone | Well-being Index for mental health tracking | Every 3 months |
| **Diet Survey** | Everyone | Nutrient intake and dietary habits (adapted from the British Heart Foundation)<sup>1</sup> | Every 3 months |
| **Physical Activity/Fitness** | Everyone | Self-reported exercise habits and motivations | Every 6 months |
| **GAD-7<sup>33</sup>** | Everyone | Generalized Anxiety | Every 6 months |
|  |  | Disorder screening |  |

Abbreviations: HRQOL - health-related quality of life

These surveys are repeated at specific intervals to track the evolution of health status over time. The use of disease-specific tools, such as the Seattle Angina Questionnaire (SAQ)^29^, Kansas City Cardiomyopathy Questionnaire (KCCQ)^30^, and EmPHasis^31^ align our smartphone application with standard outcome measures used in cardiovascular pharmaceutical and device trials, and facilitate comparisons between digital interventions and traditional clinical care.

#### Electronic Health Record (EHR) Integration

A major innovation of the next-generation *My Heart Counts* smartphone application is the integration of EHR data using HL7 FHIR standards^26^. Through HL7 FHIR, participants in the United States (and limited United Kingdom) can authorize the *My Heart Counts* smartphone application to access their electronic medical records shared through the health ecosystem (e.g., Apple Health or Google Health Connect). This will allow for automated collection of clinical laboratory values, such as low-density lipoprotein cholesterol, hemoglobin A1c, and fasting glucose levels, directly from health systems. It will also allow for identification of ICD-10 codes associated with the individual user, which can be used as clinical outcomes and correlated with self-reported survey data. For participants in the United Kingdom where this is not available, each participant will be asked to provide their National Health Service (NHS) number and consent to link their NHS data via approved services. Linkage of these traditional clinical data with data collected from the *My Heart Counts* smartphone application will allow us to robustly evaluate the prognostic value of digital biomarkers, and will also allow for retrospective linkage for pre-diagnostic assessment of historical data (where available). We note that linkage of EHR data to *My Heart Counts* is entirely voluntary and not required for participation in the larger prospective, observational cohort study or the randomized crossover trial components.

### Visualization of Cardiovascular Disease Risk

The “Heart Health” dashboard in the *My Heart Counts* smartphone application serves as a central, integrated page to summarize an individual’s cardiovascular risk. A total of 9 variables are integrated into the “Heart Health” score: weekly exercise minutes (or daily average steps for users without a smartwatch), average sleep time over the past 2 weeks, diet, mental well being, blood pressure, low-density lipoprotein cholesterol, fasting blood glucose, body mass index, and nicotine exposure (see **Table 4**). Via EHR data integration or patient self-report of traditional biomarkers, users are able to more easily understand abstract probabilities for often asymptomatic intermediate conditions (i.e., hypertension, hyperlipidemia, obesity, and diabetes). Surveys will capture an individual’s diet (via an adapted British Heart Foundation survey), mental well-being (based on the World Health Organization (WHO)-5 Well-Being Index), and nicotine exposure. Finally, via passive sensor data integration, users will be able to see how they can dynamically affect their cardiovascular disease risk over 1-2 week periods for their weekly exercise minutes or sleep time.

**Table 4.** Components of the “Heart Health” Dashboard Score.

| Variable | Data source |
| --- | --- |
| Weekly exercise minutes (or mean daily steps) | Smartwatch (minutes) or smartphone (steps) sensors |
| Mean sleep time over 2 weeks | Smartwatch, smartdevice, or smartphone sensors |
| Diet | British Heart Foundation (Survey) |
| Mental health | WHO-5 Well-Being Index (Survey) |
| Nicotine exposure | Survey |
| LDL cholesterol | Self-report |
| Blood pressure | HealthKit/Health Connect compatible device or self-report |
| Blood glucose | HealthKit/Health Connect compatible device or self-report |
| Body mass index | HealthKit/Health Connect compatible device or self-report |

Abbreviations: LDL - low-density lipoprotein; WHO - World Health Organization

### Randomization and Trial Conduct

After one week of baseline monitoring has been completed (see tasks outlined in **Table 1**), participants are provided with pre-participation information about the randomized crossover trial and given an option to opt-out if they desire^27^. Informed consent for both participation in the longitudinal cohort study and randomized crossover trial were previously presented during enrollment.

The embedded randomized crossover trial represents the study’s primary interventional objective on launch^18,19^. This trial tests whether an LLM can effectively increase physical activity levels in a free-living cohort, as compared to generic digital nudges (i.e., a reminder to reach 7,000 steps daily^34^, see **Figure 2**).

**Figure 2.**
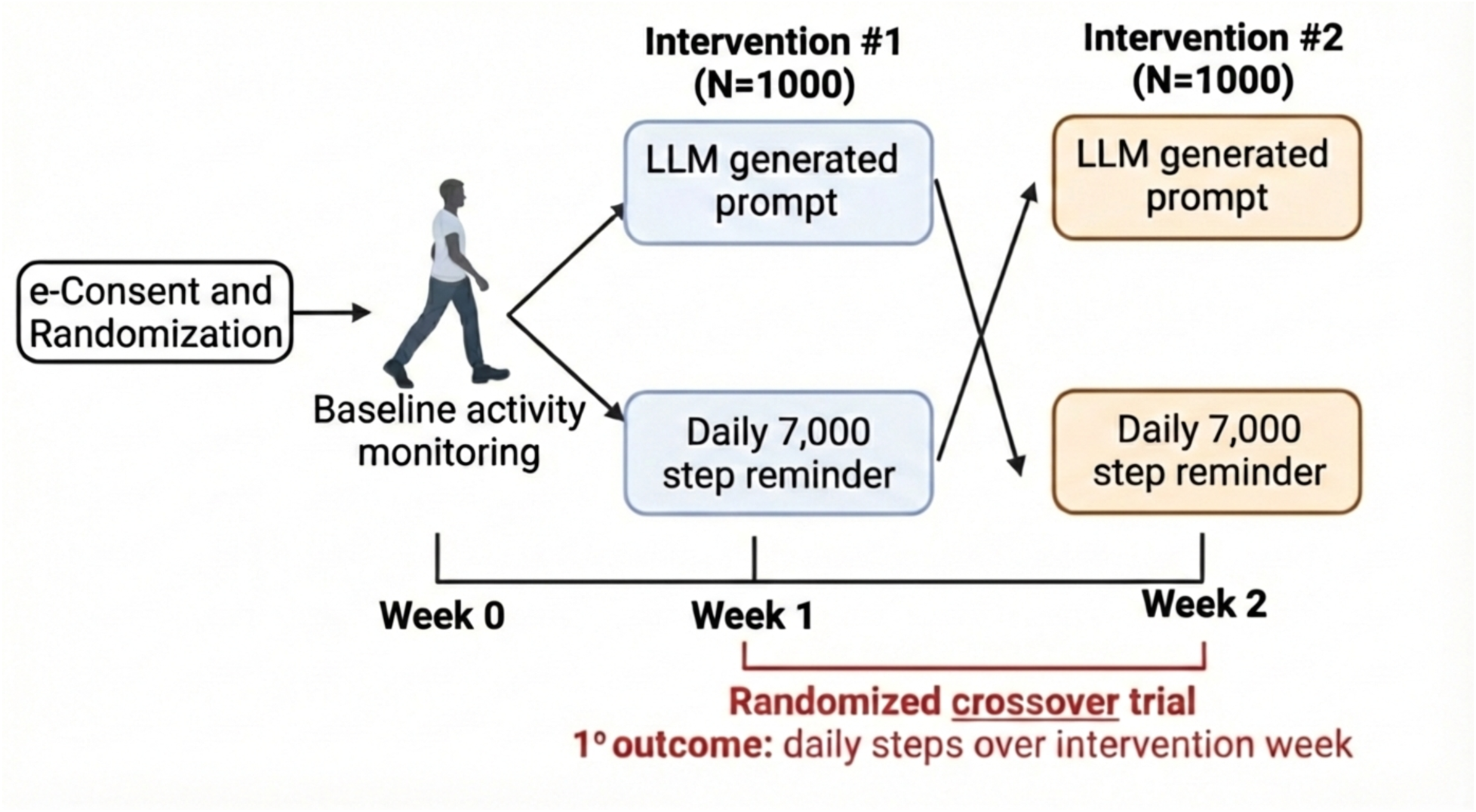
Overview of the randomized crossover trial embedded in the *My Heart Counts* smartphone application.

#### Behavioral Change Theory

The LLM used to generate personalized coaching prompts is based on the Transtheoretical Model (more commonly known as the “Stages of Change” model). This model posits that individuals transition through specific stages when adopting new health behaviors^35,36^. Prior work has found that matching personalized prompts to an individual’s specific Stage of Change yields greater responses in physical activity^35,36^. As well, prompts can evolve as a user progresses through the Stages of Change, helping to solidify long-term behavioral change.

We previously fine-tuned an LLM based on this behavioral psychology framework and found that text-based coaching prompt output was strongly preferred to those created by human expert coaches^25^. In this embedded randomized crossover trial, we will assess whether coaching prompts personalized to a user’s Stage of Change significantly increase short-term physical activity as compared to a neutral digital intervention.

#### Methodology

After consent and onboarding is complete, participants undergo simple randomization (i.e., 1:1) to receive either personalized LLM-generated text-based coaching prompts or a generic reminder to reach 7,000 steps daily, based on recent large-scale evidence^34^. They will continue to receive daily text-based notifications at their preferred time during the 2-weeks of the randomized crossover trial. After 1-week, the participants will crossover to the alternative intervention arm. During the randomized crossover trial, physical activity is passively monitored via HealthKit and Health Connect integration^18,19^. The primary outcome of interest is mean daily steps over each intervention week (LLM vs neutral control). Secondary outcomes of interest are mean active minutes (i.e., spent in moderate intensity physical activity), mean active calories burned, and the proportion of users achieving >150 minutes of moderate-vigorous activity per week for users with compatible smart-devices.

We note that while steps are the primary outcome of interest in our trial, active minutes/calories burned will likely be the focus of longer-term studies of physical activity, as once individuals find an activity they enjoy (e.g., cycling, pickleball, or swimming), their step count may not change significantly. Furthermore, we note that the *My Heart Counts* application is extensible to support longer durations for the randomized crossover trial and different distributions between intervention and control groups^22^.

This crossover design is particularly robust for digital health research, as it minimizes the impact of inter-individual variability in baseline activity, and ensures that every participant experiences the experimental intervention.

#### Statistical Power

The target enrollment of 1,000 participants provides 90% power to detect a clinically meaningful difference of 500 daily steps between the LLM coaching and neutral nudge conditions, assuming a standard deviation of 3,000 steps and accounting for 20% dropout or incomplete data. This calculation is based on a paired t-test with α = 0.05 (two-tailed) and assumes a within-subject correlation of 0.7 given the crossover design.

The 500-step threshold was selected based on previous research demonstrating that incremental increases of 500-1,000 steps per day are associated with meaningful cardiovascular benefits and are achievable for most adults^37^. Secondary power calculations indicate the study will have adequate power (>80%) to detect differences of 15-20 minutes per week in moderate-to-vigorous physical activity and clinically relevant differences in disease-specific quality of life measures for the most common cardiovascular conditions represented in the cohort.

#### Statistical Analysis

The primary analysis will employ a linear mixed-effects model to assess the effect of intervention type (LLM coaching vs. neutral nudges) on mean daily step count, accounting for the crossover design with random effects for participants and fixed effects for intervention, period, and carryover effects^18,19^. The model will adjust for baseline step count as a covariate.

The primary estimand is the difference in mean daily steps during LLM coaching periods compared to neutral nudge periods, with both interventions compared to baseline. Statistical significance will be assessed at the α = 0.05 level (two-tailed). Effect sizes will be reported as absolute differences in steps along with 95% confidence intervals and standardized mean differences (Cohen’s d).

### Feasibility

Given the modest recruitment requirement for adequate statistical power (N=1,000) and our prior recruitment (>100,000 users for the original *My Heart Counts* smartphone application)^12–19^, we believe that we will be able to complete our randomized trial as described.

## DISCUSSION

The next-generation *My Heart Counts* smartphone application and its embedded LLM coaching trial on short-term physical activity represent a significant advancement in digital health methodology, with implications for cardiovascular medicine. This release introduces several key innovations (summarized in **Figure 3)**:

**Figure 3.**
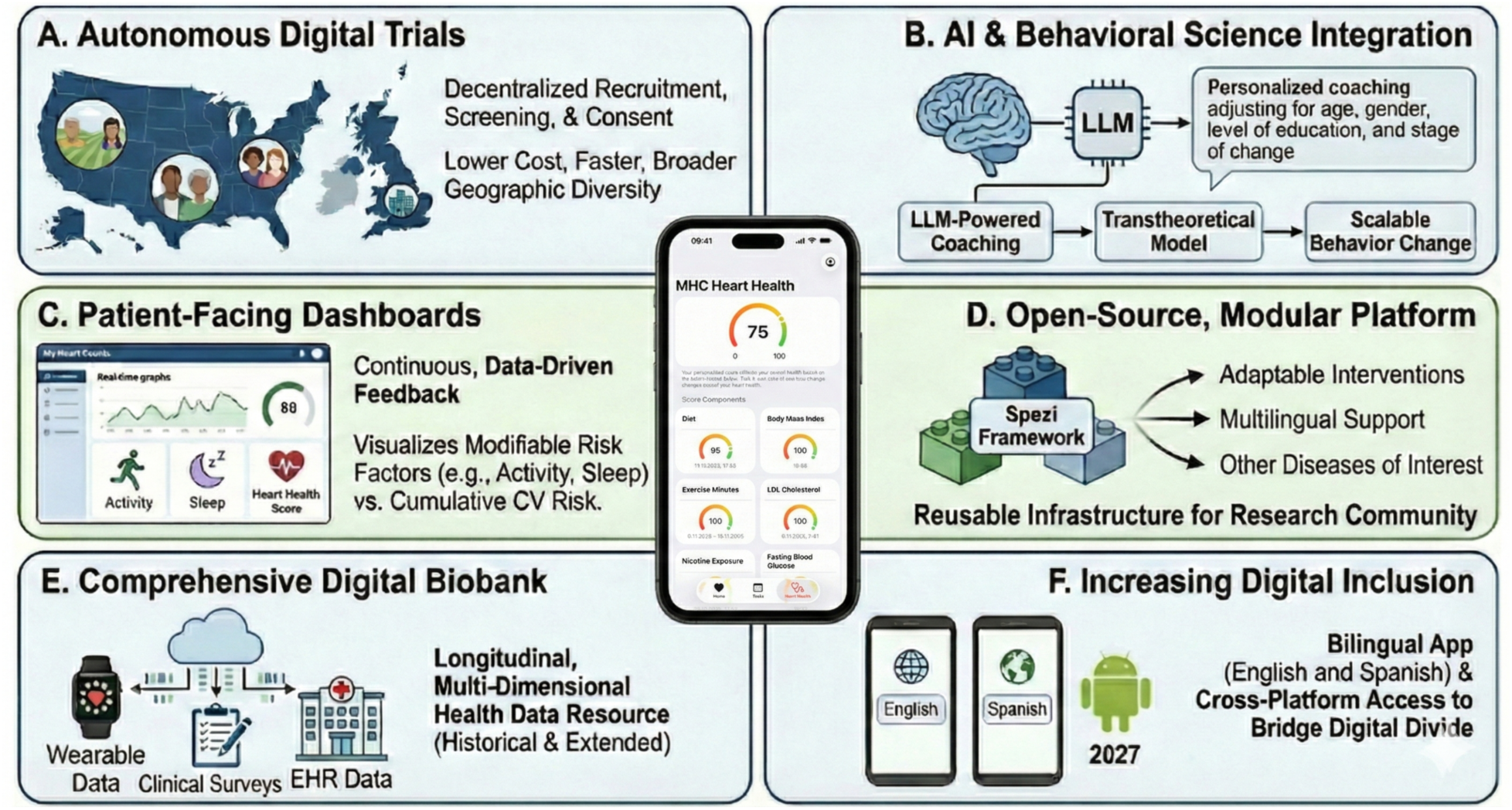
Summary of novel features in the newly-released *My Heart Counts* smartphone application. Abbreviations: AI - artificial intelligence, CV - cardiovascular, EHR - electronic health record; LLM - large language model.

### Autonomous Digital Trials in Cardiovascular Medicine

Unlike traditional clinical trials that require substantial personnel for recruitment, screening, consent, randomization, and data collection, we have previously demonstrated that selected randomized trial designs can be conducted autonomously through our smartphone application^18,19^. This autonomous approach dramatically reduces the time and cost of conducting trials while also enabling recruitment from diverse geographic regions that are often neglected in clinical trials (e.g., rural and urban areas far from academic medical centers). This elimination of geographic requirements may help address longstanding challenges with diversity and representativeness in clinical research^10,20,21,24^.

### Integration of Artificial Intelligence with Behavioral Science

The application of LLMs to generate personalized behavioral coaching interventions represents a novel synthesis of cutting-edge artificial intelligence with established behavioral psychology theoretical frameworks^25^. The Transtheoretical Model has strong empirical support, but has been challenging to implement at scale due to the personalization required^36^. LLMs offer a solution to this scalability challenge while maintaining the theoretical sophistication that drives behavioral change^25^. If successful, our approach could transform behavioral intervention delivery across multiple domains in cardiovascular health, with similar principles applied to diet modification, medication adherence, and chronic disease self-management. With the marginal cost of delivering LLM-generated interventions approaching zero at scale, the combination of LLMs with behavioral theory could make sophisticated coaching support available to populations that currently lack easy access to necessary specialists^10^.

### Patient-Facing Dashboards for Cardiovascular Risk

The incorporation of a patient-facing dashboard within the *My Heart Counts* smartphone application aims to empower users via continuous, data-driven feedback. By synthesizing objective, high-frequency wearable data with EHR-derived traditional biomarker values and patient surveys, this “Heart Health” dashboard provides a high-fidelity, real-time visualization of an individual participant’s health journey. This transition to tangible, holistic scores aims to help demystify “silent” risk factors such as hypertension and hyperlipidemia, which lack immediate symptoms, often until the onset of clinically significant disease (e.g., myocardial infarction)^6,7^. By visualizing the direct correlation between granular modifiable behaviors (such as the sensor-derived physical activity and sleep components) and cumulative cardiovascular health (along with episodic clinical biomarkers such as blood glucose, blood pressure, and lipid levels), the dashboard helps to transform static medical prognosis and probabilities into a manageable, personal goal^9^.

### Open-Source, Modular Platform

By building on the Stanford Spezi framework and releasing all code as open source, this study creates reusable infrastructure for the research community^22^. The modular software architecture can be readily modified to accommodate other studies in specific populations or with differing research questions. This approach aligns with open science principles and has the potential to accelerate digital health research across multiple domains. Some study design elements that can be potentially modified using our modular software approach are: 1) substituting different behavioral interventions into the randomized crossover trial, 2) adding disease-specific modules for conditions beyond those studied here, 3) incorporating new/additional wearable devices or sensors as they become available, and 4) implementing the platform in different languages and cultural contexts.

### Comprehensive Digital Biobank

Beyond testing a specific intervention, the *My Heart Counts* smartphone application creates a valuable resource for the research community by collecting longitudinal, multi-dimensional health data from a diverse sample across the United States and United Kingdom^13,14^. The combination of objective sensor data and ECGs (including raw data from Apple’s SensorKit), validated patient-reported outcomes (e.g., the SAQ, KCCQ, and EmPHasis surveys^29–31^), detailed demographics, and EHR data, will enable research questions that extend far beyond our primary trial and study objectives. Moreover, this biobank approach addresses a common limitation of digital health studies: short observation periods. By collecting historical data prior to enrollment into the cohort study and/or randomized trial, and also by extending data collecting beyond the initial three-week period, we can assess longer-term patterns, seasonal variations, and associations with clinical outcomes. This longitudinal dimension adds substantial value for future population health research.

### Increasing Digital Inclusion

Prior research by members of our group found that more than half of digital health trials excluded non-English speakers^21^. This both limits the generalizability of study findings and potentially worsens the digital divide, whereby those of higher socioeconomic status are more likely to benefit from advances in technology^20,21,24^. To address this, we have built parallel *My Heart Counts* smartphone applications for both English and Spanish users, as there are >17 million individuals with low English language proficiency but fluent Spanish ability currently living in the United States. We will further be developing an Android version of the *My Heart Counts* smartphone application to release in 2027 and are working on support for other languages (i.e., Mandarin Chinese and Vietnamese, among others) to further increase the digital inclusion of our cohort study and randomized crossover trial.

### Limitations and Future Directions

Selection bias represents a major limitation of digital health trials, as self-selected volunteers for smartphone-based research may differ systematically from the general population with regards to health literacy, technology comfort, and baseline health behaviors. We will attempt to mitigate this with planned development of an Android-specific smartphone application and via targeted social media advertisements to recruit demographics that are not well represented (as compared to national health survey data) in preliminary analyses to ensure generalizability to the broader population we seek to reach.

We chose the short intervention periods to minimize participant burden and maximize completion rates. However, this brief duration limits our ability to assess sustained behavior change, which is ultimately necessary for long-term reduction in cardiovascular risk (there is a short, non-sustained benefit to small increases to daily physical activity). Moreover, physical activity patterns may have a weekly or seasonal variation, which may not be fully captured with such an approach. Our future work in this space centers around the integration of just-in-time adaptive interventions and reinforcement learning, whereby the LLM learns to integrate the optimal time and place for delivery of a coaching prompt (which is further personalized to the user based on objective data and feedback)^10^.

Step count is an objective measure, but may have measurement error when considering devices (iPhone versus Apple Watch versus a variety of Android devices available). We will conduct sensitivity analyses restricted to participants with the same device types (device meta-data will be collected as part of the study to enable device-specific calibration if needed).

Separately, secondary analyses of the trial data include active minutes and active calories burned, as we appreciate that other beneficial activities (e.g., cycling, swimming, and resistance training) are not captured by step count.

### Clinical Implications

Cardiovascular disease remains the leading cause of death globally with physical activity perhaps the most critical modifiable risk factor. The *My Heart Counts* smartphone application represents a potential step toward introducing a scalable solution for promoting physical activity. By integrating artificial intelligence with behavioral psychology theoretical frameworks, personalized behavioral interventions will be accessible to the millions of individuals who could benefit from physical activity, but lack easy access to human coaching. Moreover, through parallel development in both English and Spanish, we aim to increase digital inclusion to communities and individuals that would most benefit from preventive cardiovascular care. As well, with the open-source and modular approach to software development, our smartphone application will be easily modifiable to study many conditions (e.g., Parkinson’s disease, pre-/post-surgery, cancer, etc). In addition, the visual “Heart Health” dashboard will empower participants to track and reduce their cardiovascular disease risk through small changes to their daily behaviors. Finally, with the comprehensive data sharing of our digital biobank, we aim to contribute both to immediate clinical knowledge and long-term research infrastructure in digital health.

## Data Availability

All data produced in the present work are contained in the manuscript

